# Self-tests for COVID-19: what is the evidence? A living systematic review and meta-analysis (2020-2023)

**DOI:** 10.1101/2023.08.09.23293885

**Authors:** Apoorva Anand, Fiorella Vialard, Aliasgar Esmail, Faiz Ahmad Khan, Patrick O’Byrne, Jean-Pierre Routy, Keertan Dheda, Nitika Pant Pai

**Affiliations:** Centre for Outcomes Research and Evaluation, Research Institute of the McGill University Health Centre, Montreal, Quebec, Canada; Infectious Diseases and Immunity in Global Health, Research Institute of McGill University Health Centre, Montreal, Quebec, Canada; Faculty of Medicine, McGill University, Montreal, Quebec, Canada; Centre for Lung Infection and Immunity, Division of Pulmonology, UCT Lung Institute and Department of Medicine, University of Cape Town, Cape Town, Western Cape, South Africa; Faculty of Health Sciences, University of Ottawa, Ottawa, Ontario, Canada

## Abstract

COVID-19 self-testing strategy (COVIDST) can rapidly identify symptomatic and asymptomatic SARS-CoV-2-infected individuals and their contacts, potentially reducing transmission. In this living systematic review, we evaluated the evidence for real-world COVIDST performance. Two independent reviewers searched six databases (PubMed, Embase, Web of Science, World Health Organization database, Cochrane COVID-19 registry, Europe PMC) for the period April 1^st^, 2020, to January 18^th^, 2023. Data on studies evaluating COVIDST against laboratory-based conventional testing and reported on diagnostic accuracy, feasibility, acceptability, impact, and qualitative outcomes were abstracted. Bivariate random effects meta-analyses of COVIDST accuracy were performed (n=14). Subgroup analyses (by sampling site, symptomatic/asymptomatic infection, supervised/unsupervised strategy, with/without digital supports) were conducted. Data from 70 included studies, conducted across 25 countries with a median sample size of 817 (range: 28-784,707) were pooled. Specificity was high overall, irrespective of subgroups (98.37-99.71%). Highest sensitivities were reported for: a) symptomatic individuals (73.91%, 95%CI: 68.41-78.75%; n=9), b) mid-turbinate nasal samples (77.79%, 95%CI: 56.03-90.59%; n=14), c) supervised strategy (86.67%, 95%CI: 59.64-96.62%; n=13), and d) presence of digital interventions (70.15%, 95%CI: 50.18-84.63%; n=14). Sensitivity was lower in asymptomatic populations (40.18%, 95% CI: 21.52-62.20%; n=4), due to errors in test conduct and absence of supervision or a digital support. We found no difference in COVIDST sensitivity between delta and omicron pre-dominant period. Digital supports increased confidence in COVIDST reporting and interpretation (n=16). Overall acceptability was 91.0-98.7% (n=2) with lower acceptability reported for daily self-testing (39.5-51.1%). Feasibility was 69.0-100.0% (n=5) with lower feasibility (35.9-64.6%) for serial self-testing. COVIDST decreased closures in school, workplace, and social events (n=4). COVIDST is an effective rapid screening strategy for home-, workplace- or school-based screening, for symptomatic persons, and for preventing transmission during outbreaks. This data is useful for updating COVIDST policy. Our review demonstrates that COVIDST has paved the way for the introduction of self-tests, worldwide.

## Introduction

COVID-19 cases are rapidly declining due to extensive vaccine coverage but clustering is reported in select subgroups (i.e., unvaccinated and immune suppressed individuals) [1]. A shift towards greater use of self-tests was observed towards the end of 2021. Widespread availability of rapid self-test kits, either through public distribution systems, or through private pharmacies, convenience stores, or online websites, empowered individuals to exercise autonomy in managing their exposures and guiding their actions.

COVID-19 self-testing strategies (COVIDST), are those where individuals collect their own samples, test themselves, interpret results, and use them to inform their actions post-self-test. COVIDST has particularly made the case for expanded access in the global north. However, it has greater value in areas with limited resources, in the setting of expensive/absent laboratory-based conventional testing, and in outbreaks [2].

COVIDST conducted with rapid diagnostic tests (RDTs) can detect active COVID-19 infection in a rapid turnaround time (TAT), thereby offering a convenient, user-friendly alternative to conventional reverse transcription polymerase chain reaction (RT-PCR) tests. Conventional tests require long wait times and longer TATs with increased risk of COVID-19 exposure [3, 4]. Alternatively, COVIDST can reduce dependence on healthcare workers (HCW) and reduce exposure in healthcare settings by allowing self-testing in safe, private spaces. Rapid identification of symptomatic SARS-CoV-2-infected individuals prevents exposure in community contacts and allows a timely knowledge of infection status, prompting informed action plans. Action plan initiation can greatly reduce transmission and mitigate burden on healthcare systems.

A Cochrane systematic review which assessed diagnostic accuracy of HCW-performed RDTs reported an average sensitivity of 72% in symptomatic individuals and 58% in asymptomatic individuals. However, researchers did not report outcomes beyond accuracy where RDTs were used as self-tests [5]. The World Health Organization (WHO) guidelines on COVIDST implementation released in early 2022 provide evidence on diagnostic accuracy [6]. An explosion of literature in 2022-2023 on real world performance underscores the need for a comprehensive, living review of evidence beyond diagnostic performance.

The overarching goal of this living systematic review is to update existing policies, fill evidence gaps, and provide guidance to enhance quality of tests and reporting systems in line with WHO guidelines, and to guide future outbreaks of COVID-19.

The review aims to: a) explore variability in COVIDST diagnostic performance across the spectrum of its use in a meta-analysis; b) summarize feasibility, acceptability, accessibility, and public health impact of COVIDST; and c) document qualitative outcomes.

## Methods

We registered our protocol on PROSPERO (CRD42022314799) [7]. No patients, study participants, or members of the public were involved in the design, conduct, or reporting of this review.

### Data sources and searches

Two independent reviewers (AA, FV) searched five electronic databases (Pubmed, Embase, Web of Science, WHO database, and Cochrane COVID-19 registry) from April 1^st^, 2020, to January 18^th^, 2023, for peer-reviewed journal articles and conference abstracts. Grey literature was searched through the Europe PMC pre-prints database (Fig 1). No restrictions were placed on language or publication year. We will update the review until August 1^st^, 2023.

**Fig 1.**
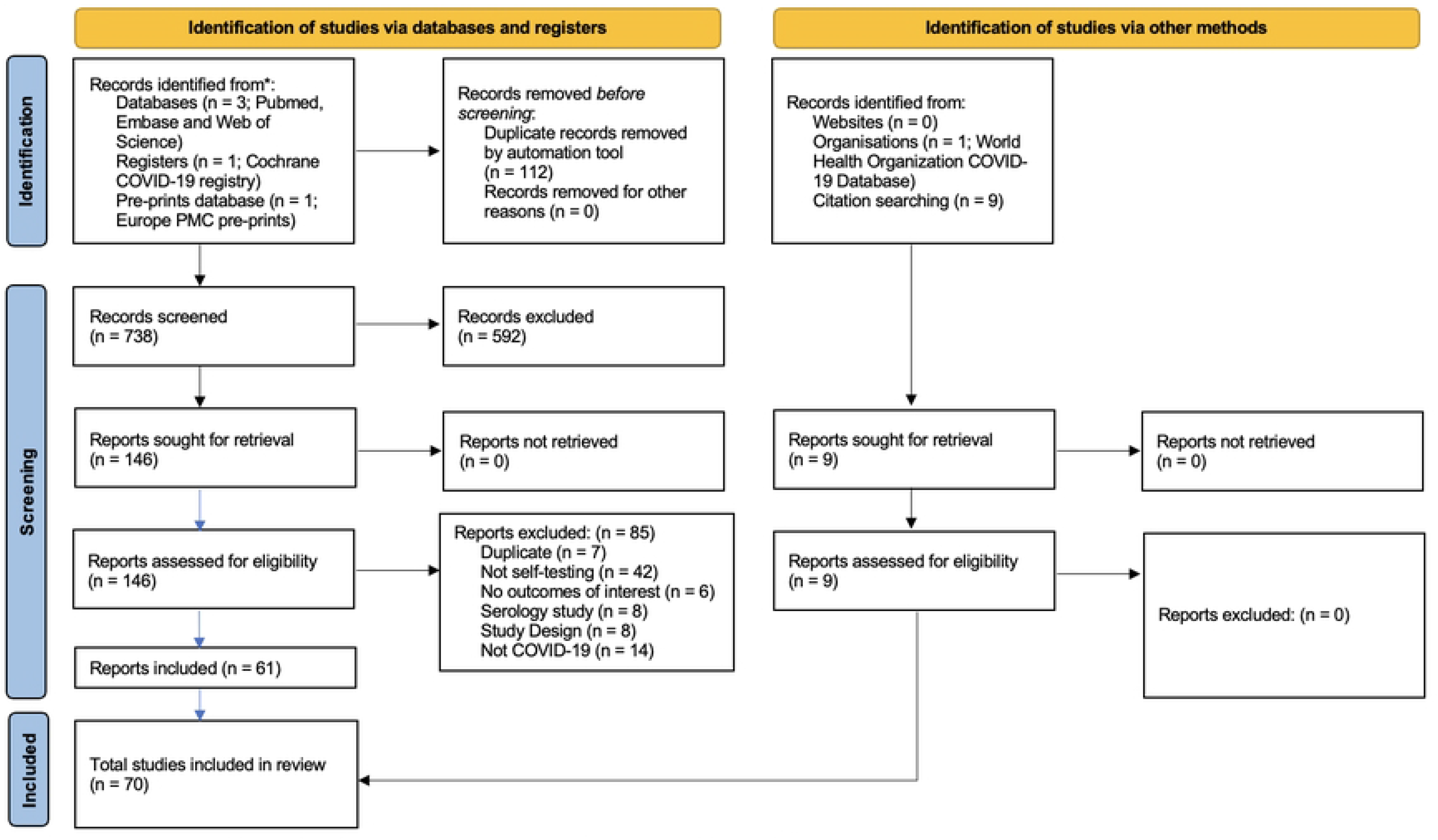
PRISMA Flow diagram

### Search string

*(COVID-19* OR covid* OR “SARS-CoV-2*”) AND (“Self-test*” OR “Self test*” OR “Self-screen*” OR “Self screen*” OR “home test*” OR “at home test*” OR “at-home test*”)* (S1 Box).

### Study selection

All studies (observational and experimental) evaluating COVIDST strategies were included. Modelling studies, commentaries, narratives, opinion pieces, review articles, and case reports were excluded. Titles, abstracts, and full texts were independently screened for eligibility based on pre-specified inclusion and exclusion criteria. Disagreements were resolved by discussion and consultation with a senior reviewer (NPP) (Fig 1).

### Data extraction and quality assessment

Data across all global geographic regions (low-, middle-, and high-income) were independently abstracted. Intervention included molecular/antigen/antibody COVID-19 self-tests. Comparators included conventional RT-PCR testing by HCWs or other trained professionals.

Primary outcome was diagnostic accuracy (i.e., sensitivity, specificity, and diagnostic odds ratio [DOR]) [8]. Authors were contacted for data when not completely available.

Secondary outcome data on feasibility, acceptability, new infections detection, preferences, and impact were abstracted and reported with summary estimates of proportions and 95% confidence intervals (CI) [8, 9]. Tertiary outcomes included qualitative measures such as motivations, facilitators, and barriers. (S1 Table).

Quality Assessment of Diagnostic Accuracy Studies Tool 2 (QUADAS-2) was used to assess risk of bias in diagnostic accuracy studies (DAS). Newcastle-Ottawa Scale (NOS) was used for observational studies and Cochrane Risk of Bias Tool 2 (RoB2) for randomized controlled trials (RCTs) [10–14].

### Data synthesis and meta-analyses

Diagnostic accuracy was explored in forest plots and heterogeneity was evaluated using *i^2^* metric. Using bivariate random-effects meta-analysis, variability in COVIDST performance across populations was explored.

Subgroup analyses were conducted for: 1) Symptom status (asymptomatic versus symptomatic individuals); 2) Strategy (supervised versus unsupervised testing strategy); 3) Site of self-sampling specimens (anterior nasal versus mid-turbinate nasal versus combined nasal-oropharyngeal versus saliva); 4) Digital support presence versus absence (i.e., websites, smartphone applications, test readers, other online tools).

All analyses were conducted in R statistical software using mada and meta packages [15, 16].

## Results

### Study selection

Seventy studies (peer-reviewed=65, preprints=5) were included. Nine of these studies were retrieved through bibliographic search (Fig 1).

### Study characteristics

Of seventy studies conducted across 25 countries, 63 (90.0%) were conducted in high-income countries (HICs) and eight (11.43%) in low- and middle-income countries (LMICs) [17]. Three studies were conducted in multiple countries [3, 18, 19]. Sample sizes ranged from 28 to 784,707 with a median sample size of 817. (S2 Table)

COVIDST strategies included mass screening (n=32), targeted screening (i.e., school, college, university, nursing home, sports club) (n=28), and healthcare facility-based screening (n=8).

Populations studied were: 1) general population members (n=39), 2) teachers, parents, school, and university students (n=11), 3) healthcare and laboratory staff (n=10), 4) hospital patients (n=5), 5) drug addiction treatment patients (n=1), 6) office employees (n=1), 7) nursing homes residents and staff (n=1), 8) music festival attendees (n=1), and 9) Black, Indigenous, and People of Colour (BIPOC) community (n=1).

Sampling sites used were anterior and mid-turbinate nasal, salivary, nasopharyngeal, and oropharyngeal.

Studies were conducted in asymptomatic (n=17), symptomatic (n=3), or both asymptomatic and symptomatic (n=27) individuals.

Thirty-four studies reported unsupervised/at-home self-testing strategy, ten studies evaluated supervised self-testing strategy, and two studies evaluated both. In supervised self-testing, the entire procedure was observed by trained HCWs or research staff, who may/may not intervene if it was incorrectly conducted or if assistance was required. In unsupervised COVIDST, unobserved testing was performed in testing centres or at-home.

Digital supports for COVIDST (n=20 studies) included websites, smartphone applications, and video-based instructions. Of these, nine studies reported digital components that aided in improving self-testing accuracy.

### Synthesized results for primary outcome (diagnostic accuracy)

Diagnostic performance of COVIDST was evaluated in two ways: A) narrative synthesis and B) meta-analysis.

First, we reported sensitivity/specificity by test devices, symptom onset, covid variants, and cycle threshold (CT) values, for which we were unable to meta-analyze due to paucity of data and studies (Narrative synthesis, Primary outcome). We reported 95% confidence intervals (95% CIs) where available. Subsequently, we generated a forest plot from pooled sensitivity and specificity (n=14). For subgroups where data was available, we conducted a meta-analysis with pooled data (Meta-analysis results, Primary outcome).

### Narrative synthesis

Diagnostic accuracy results from individual studies were summarized across test devices (n=14), by symptom onset (n=4), by CT value (n=3), and by variants (n=2). We could not perform a meta-analysis for these categories.

Four studies reported on diagnostic performance across 15 different COVIDST devices (S3 Table). Of these, four test devices reported WHO-recommended sensitivities above 80%: Boson SARS-CoV-2 antigen test card (98.18%, 95% CI: 96.74%–99.62%), Biosynex in symptomatic populations (93.8%; 95% CI: 79.3%–98.4%), Biosynex in asymptomatic populations (83.3%; 95% CI: 73.4%-90.0%), Standard Q by SD Biosensor (82.50%, 95% CI: 68.1%–91.3% and 94.38%, 95% CI: 87.54%-98.60%), and MP Bio (83.01%, 95% CI: 78.8%-86.7%). Specificities were above 91% for all devices. [20–26].

Four studies reported accuracies by day of symptom onset. Two studies reported sensitivities of 99.18% one day prior to symptom onset, 98.77-100% on first 2 days, and 100% from day 2 to 7 of symptom onset [20, 25]. Conversely, a community-based study reported sensitivities of 23% within 0-1 days and 66.67% within 2-4 days of symptom onset [23]. Finally, another study reported a sensitivity of 73% when self-test was conducted within 0-5 days of symptom onset as compared to 22% when conducted after 5 days [27].

Three studies reported on self-test performance by cycle threshold (CT) values. Low CT values of positive RT-PCR results indicated a high viral load in swab samples. RT-PCR and self-test results were compared; CT value was checked for each self-test result. One study detected 100% of infections with COVIDST when CT values were below 20, 92% when CT values were between 20-30, and 33.33% when CT values were above 30 [20]. COVIDST in 2 studies detected: 1) symptomatic cases when mean CT value was 23.1 (IQR: 19.5–30.0) and median CT value was 14 (IQR: 12.0-18.0); 2) asymptomatic cases when mean CT value was 28.2 (IQR: 25.0–33.0) [23, 28].

Two studies compared COVIDST performance in delta versus omicron variant infected populations. In one study, sensitivities decreased from 87.0% in the delta period to 80.9% in the omicron period [26]. Conversely, in another study, same-day sensitivity of self-tests was higher (22.1%, 95%CI: 15.5-28.8%) in omicron period versus 15.5% (95%CI: 6.2-24.8%) in delta period [29].

### Meta-analyses results

Fourteen studies reported data on accuracy [19–26, 30–35]. First, we pooled sensitivities and specificities to create forest plots (S1 Fig A and B). Following this, we assessed heterogeneity and conducted subgroup analyses; results are summarized below.

Our forest plots reported a point estimate for pooled sensitivity (n=14) of 75.0% (95%CI: 59.0%-86.0%) (S1 Fig A). Sensitivities varied from 25% to 98%. Random effects model heterogeneity *i^2^* statistic was high at 97%. Point estimate for pooled specificity (n=14) was 100% (95%CI: 99.0%-100.0%) (S1 Fig B). Specificities varied from 97% to 100%. Random effects model heterogeneity *i^2^* statistic was high at 94%.

We conducted subgroup analyses to explore this heterogeneity further. Summary receiver operating characteristic (SROC) curves were plotted for all subgroups (S2 Fig A-D). Pooled sensitivities, specificities, and DORs estimates are provided in Table 1.

**Table 1.** Meta-analyses of COVIDST diagnostic accuracy.

| Sr No | Category | Sub-groups | Sensitivity | 95% CI | Specificity | 95% CI | DOR | 95% CI |
| --- | --- | --- | --- | --- | --- | --- | --- | --- |
| 1 | Specific sampling site | Anterior nasal | 63.8 | 46.68-78.00 | 99.29 | 98.73-99.60 | 263 | 123-497 |
|  |  | Mid-turbinate nasal | 77.79 | 56.03-90.59 | 98.62 | 94.35-99.67 | 291 | 84.1-741 |
|  |  | Saliva | 39.1 | 18.45-64.57 | 99.32 | 97.60-99.81 | 98.8 | 55.50-163.00 |
|  |  | Combined nasal-oropharyngeal | 69.69 | 58.96-78.62 | 99.18 | 97.84-99.69 | 303 | 124-625 |
| 2 | Symptomatic status | Symptomatic | 73.91 | 68.41-78.75 | 98.37 | 97.47-98.95 | 175 | 108-270 |
|  |  | Asymptomatic | 40.18 | 21.52-62.20 | 99.71 | 99.29-99.88 | 249 | 104-508 |
| 3 | Testing method | Supervised | 86.67 | 59.64-96.62 | 99.39 | 97.04-99.88 | 1530 | 200-5670 |
|  |  | Unsupervised | 60.69 | 50.31-70.18 | 99.13 | 98.60-99.47 | 181 | 116-269 |
| 4 | Digital Intervention | Present | 70.15 | 50.08-84.63 | 99.39 | 97.99-99.82 | 409 | 193.00-764.00 |
|  |  | Absent | 65.69 | 54.06-75.70 | 99.17 | 98.66-99.49 | 237 | 135-387 |

| Significance code | P-value |
| --- | --- |
| *** | [0, 0.001) |
| ** | (0.001, 0.01) |
| * | (0.01, 0.05) |
| . | (0.05, 0.1) |
|  | (0.1, 1) |
| Reference variable |  |

In subgroup analyses by sampling sites (n=14), highest sensitivity was reported in samples from mid-turbinate sampling (77.79%, 95%CI: 56.03%-90.59%), followed by combined nasal-oropharyngeal sampling (69.69%, 95%CI: 58.96%-78.62%), and anterior nasal sampling (63.80%, 95%CI: 46.68%-78.0%, statistically significant). Sensitivity was lowest with salivary sampling (39.10%, 95%CI: 18.45%-64.57%). Specificity was above 98% irrespective of sampling site. DOR was highest for combined nasal-oropharyngeal specimens (303.00) and lowest for saliva specimens (98.80).

Nine out of fourteen studies reported diagnostic accuracy data based on presence/absence of symptoms. For symptomatic populations, sensitivity was 73.91% (95%CI: 68.41%-78.75%, statistically significant) versus 40.18% (95%CI: 21.52%-62.20%) for asymptomatic populations. Specificity was above 97% irrespective of symptomatic status. DOR was high at 249 for asymptomatic versus 175 in symptomatic populations.

Thirteen out of fourteen studies evaluated performance of supervised and unsupervised COVIDST. Supervised strategy reported a higher sensitivity of 86.67% (95%CI: 59.64%-96.62%) versus a sensitivity of 60.69% (95%CI: 50.31%-70.18%) in unsupervised strategy. Specificity was high at 99% irrespective of strategy. DOR was higher in supervised (1530.00) versus in unsupervised (181.00) COVIDST.

Fourteen studies analyzed COVIDST performance with/without digital supports. Sensitivity was higher with digital supports (70.15%, 95%CI: 50.08%-84.63%) than without (65.69%, 95%CI: 54.06%-75.70%). Specificity was 99% irrespective of presence/absence of digital supports. DOR was higher (409.00) with digital supports than without them (237.00).

### Synthesized results for secondary outcomes

#### Test positivity (new infections detected)

Across twenty studies, new infections detected by COVIDST varied from 0.02% to 27% [22, 25, 27, 28, 31, 36–48]. In two other studies, test positivity varied from 12% to 83.3% during the delta wave and 41.7% to 87.2% during the omicron wave [29, 49]. Point prevalence for at-home COVIDST was 3.7% compared to 5.5% for testing by HCWs .[47].

#### Acceptability and willingness to use

Thirteen studies reported an overall high acceptability and willingness to use COVIDST. *C*OVIDST acceptability was high (91%-98.7%) in two studies, with higher acceptability in females (73.91%) versus males (60.09%) reported in another study [50–52]. Acceptability was lower (39.48%-51.1%) for daily self-testing [38, 40, 52]. Hesitancy to test (33.8%) and concerns about test accuracy (1%) made people decline COVIDST [40].

Across three studies in different populations, COVIDST uptake was 97% in school children, 92.5% in children with medical problems, and 45.2% in a mass self-testing study [41, 43, 53]. Across seven studies, willingness to use nasal self-tests ranged from 77% to 95.8% [2, 54–59].

#### Feasibility and usability

Eighteen studies reported high COVIDST feasibility and ease of use. Usability threshold, defined as the ability to correctly conduct all critical self-test steps, was higher with digital supports.

An overall high feasibility was reported (69.6%-100%) across five studies [23, 40, 45, 60, 61]. In three studies, feasibility was lower for serial-testing COVIDST (35.9%-64.6%) [41, 50, 62]. The average completion rate was 4.3 self-tests over 4.8 weeks in another serial-testing study [62].

Across seven studies, participants found COVIDST easy to use (81%-100%) [22, 30, 34, 45, 59, 63, 64]. Specifically, two studies reported a high ease of conducting at-home self-tests (95.7%), ease of reading self-test results (92%), and ease of remembering to test regularly (96%) [22, 38].

Across four studies, confidence in reporting test results and testing abilities was high (70%-98%) [30, 34, 38, 65]. Regular COVIDST by dentists improved perception of safety while treating patients by 49% [66].

Usability threshold was assessed in three studies. A high usability threshold was reported from Malawi (82.4%-90.4%) and Zimbabwe (65.4%-70.6%) [2]. In Germany, usability was 61.2%, while in France, it increased from 99.1% to 100% with video supports [23, 67].

#### Preference

Across six studies, preference for COVIDST varied from 29% to 87.9% [32, 45, 51, 64, 65, 67]. Overall, COVIDST preference was higher among white people, urban populations, political right, individuals with a college degree, and healthcare workers, as compared to ethnic minorities, rural populations, political left, individuals with a lower education, and working in other occupations [32, 51, 59, 63, 68, 69]. 94% of participants preferred throat swab-based self-test and 90% preferred saliva-based self-tests [55]. In another study, 95.4% participants preferred over-the-counter vending machines to obtain self-test kits [70].

#### Impact outcomes

Impact outcomes were evaluated in eighteen studies. In four studies, COVIDST reduced closures in different institutions and of public events. Regular COVIDST in a peri-urban primary school resulted in fewer school closures and decreased secondary infections in one study [71]. In another, daily mass COVIDST resulted in 8,292 workday savings of essential workers [41].

Self-tests were also used as daily testing tools in high exposure HCWs, allowing them to quarantine immediately in case of a positive result and prevent infection transmission [28]. In addition to healthcare settings, COVIDST facilitated the safe working of co-working health laboratory sites in a pandemic setting [31]. Furthermore, pre-event COVIDST allowed attendees to safely enjoy music concerts, wherein 87% of self-testers perceived a lower risk of contracting COVID-19 at the concert [72].

Three studies reported a higher TAT with COVIDST compared to conventional testing. In one study, TAT of 15-30 minutes for COVIDST versus 24-48 hours for RT-PCR was reported [22]. Antigen self-tests had a mean TAT of 8.1 minutes (standard deviation: 1.3) [23]. In another study, self-tests identified 23.5% of infections within 24 hours, and 54.9% of infections in the next 48 hours, prior to obtaining RT-PCR results.

Impact of COVIDST on action plans (n=7) and self-test result notification (n=4) was reported. In four studies, willingness to notify close contacts and relevant authorities was 80%-97.6% [2, 52, 54, 57]. In two studies, a high proportion of respondents (80.78%-98.32%) were willing to seek post-test counselling following a positive result [52, 57]. In three studies, 93%-100% testers expressed willingness to self-isolate following a positive test result [2, 57, 73]. Although only 49% of HCWs believed that self-testers would self-isolate themselves following a positive result, they opined that self-testers would take steps to reduce infection transmission [2].

Across two studies, 54%-78.3% of participants preferred validating initial COVIDST results through repeat testing [23, 54, 57]. In three studies, 70.1%-92.6% self-testers sought confirmatory RT-PCR testing [39, 41, 54]. Children aged 5-11 years and 12-18 years with a positive unsupervised self-test result were more likely to obtain a confirmatory PCR test compared to supervised testers (Odds ratio=3.48, 95%CI: 2.68-4.52 and Odds ratio=2.16, 95%CI: 1.86-2.50, respectively) [36].

#### Qualitative outcomes

Qualitative outcomes such as motivations, facilitators, and barriers were assessed in 26 studies.

Motivators to self-test were protecting one’s health and reducing infection transmission to close contacts, partaking in daily activities and physically accessing services, workplace safety, travelling, dining outside, and attending large gatherings [45, 54, 67, 72, 74, 75]. Higher motivations to test were linked to a higher socioeconomic status (SES) and ability to acquire test kits [45, 69, 72, 74].

COVIDST facilitators assessed in twelve studies included self-test training prior to use, non-intrusive and ease of testing at-home, increased sense of safety, detailed self-test instructions, faster turnaround time, and instructional videos [23, 28, 32, 57, 61, 62, 67, 69, 71, 76–80].

Across nine studies, COVIDST barriers included high costs, low trust in accuracy and reliability, anxiety, hesitation in self-test conduct, uncomfortable self-swabbing procedures, difficulty following instructions and interpreting faint positive test lines, lack of perceived benefit, and inequitable access to COVIDST [21–23, 53, 62, 79, 81–83].

#### Self-testing with digital supports

Across fifteen studies, COVIDST digital supports used were: online platforms (n=6), app-based COVIDST (n=6), video-based instructions (n=5), and online supervised COVIDST (n=6) [22–24, 32, 34, 38–40, 45, 46, 49–51, 59–62, 65, 72, 84].

In four studies, app-assisted COVIDST allowed 98%-100% of participants to successfully interpret their test results,[38, 50, 60, 84]. while video-taped self-testing process increased participants’ confidence (76%) in COVIDST results [72].

In another four studies, uploading a test result picture or reporting test results online was a requirement that allowed HCWs to monitor and isolate positive cases [40, 49, 51, 60, 65].

In a mass COVIDST study, digital supports increased result notification in 75% of self-testers [84]. A self-testing and COVID-19 exposure notification app utilized such self-reported COVIDST results to reduce risk of infection in non-infected app users. [48, 49]. However, unincentivized and voluntary reporting with a digital assistant in one mass COVIDST study was low (4.6%) [84]. Also, digital reporting varied by test result; 3.2% reported positive test results and 1.8% reported negative test results [60]. One study reported that federal COVID-19 statistics did not include 42.8% of participants with a positive self-test result [48].

### Risk of bias assessment

To assess any publication bias in studies included in the meta-analysis, a funnel plot was plotted (S3 Fig). A low risk of bias was estimated using Deek’s method (p-value of 0.79).

Using the QUADAS-2 tool (n=14), we found low risk of bias across all categories except for reference standards (unclear risk, n=6) (S4 Table A). Cohort studies (n=13) had an average risk of bias in the comparability category (1-star, n=7) (S4 Table B). Similarly, cross-sectional studies (n=41) also had an average risk of bias in the comparability category (1-stars, n=12) (S4 Table C). One case-control study had an overall poor risk of bias score across all categories. Finally, RoB2 tool was used for one qualitative RCT study wherein low risk for all domains was observed except for the selection of reported result domain.

## Discussion

This review demonstrates that COVIDST strategies are effective in screening SARS-CoV-2 infections. Self-testing has a faster TAT compared to conventional testing, can be used in outbreak settings, prevent institutional closures, and reduce infection transmission in various settings.

### Diagnostic accuracy and caveats

Our meta-analyses demonstrated very high specificity and above average sensitivity of COVIDST strategies.

Specificity for COVIDST (across all tests) was consistently above 98% regardless of different subgroups. Specificity is computed by calculating all true negatives (TN)/true negatives (TN) and false positives (FP). If the specificity is high, and the person is asymptomatic, we can be certain that the false positives are low.

In contrast, sensitivity for COVIDST varied across subgroups; highest sensitivities were reported for: a) mid-turbinate nasal specimens (77.79%, 95% CI: 56.03%-90.59%), b) tests conduct in supervised settings (86.67%, 95% CI: 59.64%-96.62%), c) symptomatic individuals (73.91%, 95% CI: 68.41%-78.75%), and d) digital COVIDST (70.15%, 95% CI: 50.08%-84.63%).

Sensitivity is computed by reporting true positives (TP)/true positives (TP) plus false negatives (FN). With that, if false negatives increase, sensitivity falls. In symptomatic individuals, highest sensitivities were reported for the first 5 days of symptom onset. In contrast, for asymptomatic individuals, sensitivities were consistently low (40.18%). Interpreting sensitivity and specificity is tricky at the population level, therefore simple messaging is very important for populations seeking to implement or use these self-tests. Our results show lower COVIDST sensitivity were due to unclear instructions for use, inadequate pre-test training, incorrect test conduct, non-adherence to instructions, and difficulties in interpreting faint positive test lines. To improve COVIDST performance, diagnostic companies need to design self-test kits with consideration for low-literacy and senior populations. Self-test instructions for conduct and interpretation must be detailed, comprehensive, and provided in layman terms. In areas with high digital literacy and data connectivity, video-based instructions and virtual pre-test training sessions can be provided.

Additionally, sensitivities were higher when CT value was lower or equal to 25. This is an important feature to note while sharing information on self-tests. Variance in sensitivity based on CT values show that self-tests can detect infections most accurately with peak viral loads and contagiousness. These findings highlight that the value of self-testing lies in the rapid identification and prompt isolation of highly contagious individuals compared to RT-PCR positive tests. A median PCR positivity period of 22-33 days gives a positive test result in the presence of viral particles that persist even after resolution of infection.

Comparatively, most false negative self-test results occur when individuals are outside the transmissibility window [85]. If a COVIDST result is negative but an RT-PCR test result is positive, it is likely that the individual is not very infectious and may not pose a public health threat [85].

As for test devices used, some devices performed consistently as per WHO – for example, the Boson SARS-CoV-2 antigen test card, Biosynex, Standard Q at-home test, and MP Bio – while others did not (Please refer to Appendix table 3). Regarding strains, in the two studies that evaluated COVIDST performance by variants, we noted no difference in sensitivities for either the delta or the omicron strain pre-dominant periods. This is reassuring for future strains of the virus.

### Secondary and tertiary outcomes

COVIDST screening strategies offer benefits in pandemic settings, when accessibility to laboratory testing is very limited, and timely test results are of the essence. Our results show that COVIDST strategies consistently reported a rapid TAT, were overall highly acceptable, highly feasible, and convenient to populations around the world. Their usability index was at 100% with additional digital supports. These supports included video-based or app-based instructions, highlighting the potential of digital COVIDST.

Our results are consistent with the interim guidance on self-testing provided by WHO, which found self-testing acceptable, feasible, and easy to use by laymen; however, our results are updated and include data that can serve WHO to adapt their guidance. These results were also very similar to the proven benefits that have been demonstrated with HIV self-testing [86].

Despite established COVID-19 nucleic acid amplification testing (NAAT) surveillance systems in many countries, COVIDST became an important screening and decision-making tool for individuals during the peak of the pandemic [87]. Our results show that regular COVIDST was instrumental in impacting onward transmission that stemmed from the pandemic. This impact was demonstrated in reducing school closures, resuming in-person education, and allowed attendees to safely attend social events. Healthcare workers were able to treat patients while monitoring themselves, thereby reducing the risk of nosocomial infections.

Serial testing during the pandemic, especially in high exposure jobs, allowed essential workers to resume work without fear of losing pay and laboratories were able to remain operational. Participants were willing to report results, adhere to self-isolation guidelines, and seek confirmatory testing following a positive self-test result. Periodic self-testing reduced anxiety and created an environment of safety and reassurance when resuming normal activities.

Overall, participants were motivated to use COVIDST strategies to know their infection status, resume daily activities, protect their loved ones, and exercise caution while attending large gatherings. Motivations and preference for COVIDST over lab-based testing increased with a higher SES and in urban areas.

Inequitable access to self-tests in ethnic minorities with a lower SES was observed. This alludes to inequity in distribution of self-tests that was largely restricted to those with resources. This pattern could be changed for future pandemics by reducing the unit price of self-tests and public procurement of tests for large scale use.

Evidence on COVIDST parallels the vast evidence that has accumulated for HIVST. Both viruses have paved the way for a greater use of self-testing solutions to know serostatus, and by increasing accessibility offered by these solutions during the pandemic, have made self-tests a common household name. This strategy holds promise for many infectious pathogens and pandemics.

### Strengths and limitations

To our knowledge, ours is the first comprehensive and updated systematic review and meta-analysis on COVIDST. Although WHO released COVIDST guidelines, data on diagnostic accuracy then were scarce, so meta-analyses could not be performed. Additionally, these guidelines were based on studies published before February 2022 while our updated review contains recent studies (2023) that complement these results.

Our review is based on observational data. With a few RCTs underway, new data will soon become available [88, 89]. Most of the data are from HICs (n=63), making our results difficult to generalize to LMICs. Finally, no studies were reported with highly accurate molecular rapid COVIDST strategies [90].

### Implication for product development and research

Publicly distributed self-tests can guarantee widespread accessibility but should be implemented with evidence-based strategies to improve test conduct and result interpretation. Checks for counterfeit test kits are necessary and regulating the sales of COVIDST kits can help improve public confidence in self-testing.

Public health sector and not-for-profit organizations along with healthcare facilities and pharmacies can increase access to self-tests by free-of-cost, widespread distribution of kits in urban and rural areas.

A strong and connected reporting system must be implemented by local authorities to avoid underestimating the true burden of infections. Future research can explore COVIDST diagnostic performance with digitally connected platforms, apps, test readers, and systems to report message notification and linkage to care. Data from clinical trials are needed to fill the gaps in evidence from LMICs.

## Conclusion

Self-testing complements conventional testing in the pandemic setting with its speed and efficiency when time is of the essence. Our review demonstrates that COVIDST is a convenient and effective strategy for screening infections when used by the general population.

In symptomatic populations, in supervised settings with guided instructions, and with the addition of digital supports, self-tests improved in their performance. COVIDST had a high usability threshold, impacted institutional closures, and reported results notification where reporting systems were in place. However, data from LMICs were limited due to scarcity of self-testing.

Digital COVIDST is promising, and additional data will help improve accuracy and trust. Our results can aid policymakers, government bodies, and healthcare systems in updating their policies, and organizations aimed at integrating triple self-testing strategies in their health ecosystems. COVIDST can alleviate the impact of the COVID-19 pandemic across all global settings and their widespread availability will help address global health inequities.

Both HIVST and COVIDST have demonstrated the impact that self-tests can have in empowering lay individuals to know their serostatus and in preventing forward transmission. This approach holds promise for the many self-tests for related pathogens (HCV, HBV, Syphilis, and MPox), and use of this tool can aid in ending future waves of these pandemics.

## Acknowledgements

The authors would like to thank Olivia Vaikla for her assistance with proofreading and formatting the manuscript.

## Authors’ contributions

AA and NPP designed, drafted, and reviewed the initial manuscript while the remaining authors (FV, FK, AE, POB, JPR, KD) provided critique on subsequent drafts. Search strategy was developed and executed by AA, FV, and NPP. Data abstraction and quality assessment were performed by AA and FV. All authors (AA, FV, FK, AE, POB, JPR, KD, NPP) have reviewed and approved the final written manuscript.

## Data availability

All data relevant to the study are included in the article or uploaded as supplementary information.

## Supporting information

**S1 Box. Search String.**

**S1 Table. Outcomes Description.**

**S2 Table. Study Characteristics.**

**S3 Table. Diagnostic accuracy across test devices.**

**S1 Fig A and B. Forest Plot.**

**S2 Fig A-D. Comparison of summary receiver operating characteristic (SROC) curves.**

**S3 Fig. Funnel Plot of studies included in the meta-analysis (n=14).**

**S4 Table A-C. Risk of bias.**

